# Teacher-rated aggression and co-occurring behaviors and problems among schoolchildren: A comparison of four population-based European cohorts

**DOI:** 10.1101/19002576

**Authors:** A Whipp, E Vuoksimaa, K Bolhuis, EL de Zeeuw, T Korhonen, M Mauri, L Pulkkinen, K Rimfeld, RJ Rose, T CEM van Beijsterveldt, M Bartels, R Plomin, H Tiemeier, J Kaprio, DI Boomsma

## Abstract

Aggressive behavior in school is an ongoing concern, with the current focus mostly on specific manifestations such as bullying and extreme violence. Children spend a substantial amount of time in school, but their behaviors in the school setting tend to be less well characterized than in the home setting. Since aggression may index multiple behavioral problems, we assessed associations of teacher-rated aggressive behavior with co-occurring externalizing/internalizing problems and social behavior in 39,936 schoolchildren from 4 population-based cohorts from Finland, the Netherlands, and the UK. Mean levels of aggressive behavior differed significantly by gender. Correlations of aggressive behavior were high with all other externalizing problems (0.47-0.80) and lower with internalizing problems (0.02-0.39). A negative association was seen with prosocial behavior (−0.33 to −0.54). Despite the higher mean levels of aggressive behavior in boys, the correlations were notably similar for boys and girls (e.g., aggressive-hyperactivity correlations: 0.51-0.75 boys, 0.47-0.70 girls) and did not vary greatly with respect to age, instrument or cohort. Thus, aggressive behavior at school rarely occurs in isolation and children with problems of aggressive behavior likely require help with other behavioral and emotional problems. It is important to note that greater aggressive behavior is not only associated with greater amount of other externalizing and internalizing problems but also with lower levels of prosocial behavior.

## Introduction

Aggressive behavior in school is a persistent topic of concern, with bullying, gender- and sexuality-based misconduct, and extreme violence (e.g., school fights, stabbings, shootings) currently garnering the most attention (OECD, 2017; United Nations Educational, 2019). While these specific manifestations of aggression are important to understand and manage for the safety of all in the school environment, aggression may be a marker for a wider set of behavioral problems that co-occur with it. A more complete picture of the problems of aggressive behavior in school is achieved when we also understand the general levels of the behavior and the context within which the behavior occurs Aggression is a heterogenous behavior that has been characterized in a variety of ways, including well-researched subtypes such as proactive (i.e., planned aggression, such as bullying or stealing), reactive (i.e., in response to some stressor, such as a bully instigating a confrontation), direct (i.e., yelling, hitting, kicking) and indirect (i.e., social/relational aggression, such as spreading rumors or ostracizing an individual) (Pulkkinen, 2018; Vitaro, Brendgen, & Barker, 2006). All children express such behaviors to varying degrees, in time and amount. This complex behavior among school-aged children is associated with relationship quality among their peers, teachers, and families, as well as their eductional attainment, risk for substance abuse and other psychiatric disorders, and risk for partipation in criminal activities (Farrington, 1995; Pulkkinen, 2017; Whipp et al., 2019).

In studying aggression, gender differences have played a large and complex role in our understanding of the problem behavior. Early research focused only or mainly on boys, although studies in recent decades have greatly enriched our knowledge regarding both genders. Distinctions between direct/physical aggression as being higher among boys and indirect aggression being higher among girls have generally given way to an understanding that both genders engage in both types of aggression, and that at least for indirect aggression the gender differences, if any, are small (Card, Stucky, Sawalani, & Little, 2008; Cleverley, Szatmari, Vaillancourt, Boyle, & Lipman, 2012). Furthermore, a study by Nivette et al. (2018) indicates that in societies that experience high gender inequality, differences in physical aggression by gender are minimal.

Complicating our understanding of this behavioral problem further, children with aggressive behavior often have co-occurring problem behaviors, either externalizing, internalizing or both (August, Realmuto, Hektner, & Bloomquist, 2001; Bartels et al., 2018; Verhulst & van der Ende, 1993). Furthermore, children with aggression and co-morbidities often have more than one co-morbidity, worse prognoses and poorer outcomes (Fontaine et al., 2008; Pulkkinen, 2017; Verhulst & van der Ende, 1993). Regarding the school setting, research has generally focused on severe aggression diagnoses (e.g., conduct disorder, oppostional-defiant disorder), with less attention on general, non-pathological levels of aggressive behavior and co- occuring behaviors. To shed light on the overall phenotype of aggressive behavior in schools and how common co-occuring behaviors generally are, we carried out a unique study in four large population-based cohorts that assembled ratings from teachers of children.

Aggressive behavior is often characterized and studied via parental, self, and mental health professional assessments, with teacher ratings being available less often in epidemiologic or clinical research. Research from the past several decades has shown that an individual’s aggressive behavior may be situational (e.g., present at school, but not at home) and that assessments of behavioral and emotional problems do not correlate well between raters (Achenbach, McConaughy, & Howell, 1987; De Los Reyes & Kazdin, 2005; Verhulst, Koot, & Van der Ende, 1994). Furthermore, studies have shown that teacher ratings of problem behaviors have often been most useful in diagnostics and predictive outcomes, including psychiatric disorders and criminality (Carbonneau, Tremblay, Vitaro, & Saucier, 2005; Hodgins, Larm, Ellenbogen, Vitaro, & Tremblay, 2013; Verhulst et al., 1994; Whipp et al., 2019). Teachers’ ability to observe children in a complex task-based setting and among peers of similar ages and abilities provides them with a valuable comparison-base, making their insight unique and important.

This study’s overall objective was to characterize the levels and associations of aggressive behavior and co-occurring behaviors in the school setting, using teacher ratings of children ages 7–14. Large datasets from collaborating population-based cohorts of children from Finland, the Netherlands, and the UK were analyzed. Specific aims were: 1) to report the mean levels of aggressive behavior and other behaviors and emotional problems as assessed by teachers, 2) to examine associations (co-occurrence) of aggressive behavior and other behaviors and emotional problems, and 3) to assess gender differences in the mean levels and associations of aggressive behavior and co-occurring behaviors and emotional problems.

## Materials and Methods

The datasets for this investigation were obtained through the collaboration of the ACTION (Aggression in Children: Unravelling gene-environment interplay to inform Treatment and InterventiON strategies; http://www.action-euproject.eu/) consortium (Bartels et al., 2018). Of the seven large child/adolescent cohorts brought together in ACTION, four had collected teacher rating data. In total, the data included 39,936 teacher ratings on children at ages 7, 9, 10, 12, and 14 (48.9% boys) from Finland, the Netherlands, and the UK, with some children observed at two or more ages. As in Bartels et al. (2018), in the cohorts that included twin data one twin per family was randomly selected for analysis. Individuals were excluded if they had an illness or handicap that interfered with their daily functioning, and if they were missing substantial data from one of the examined subscales from cohort-respective behavioral and emotional problem questionnaires (see also: http://www.action-euproject.eu/content/data-protocols). All data in the current investigation were collected under protocols that have been approved by the appropriate ethics committees, and studies were performed in accordance with the ethical standards established in the 1964 Declaration of Helsinki and its later amendments. Brief cohort and behavioral questionnaire descriptions are presented here (see Online Resource 1 for further details on teacher rating collections and the school systems in Finland, the Netherlands, and the UK).

### Cohort descriptions

The FinnTwin12 (FT12) dataset was established in Finland from the cohort of twins born 1983–87 (Kaprio, 2013). It is a population-based twin study aimed at examining health-related behaviors and their precursors. Data were collected at ages 12, 14, 17, and 22. Teacher ratings on behavioral and emotional problems were collected at ages 12 and 14. Response rates were 93% and 94% for age 12 and 14 teacher ratings, respectively.

The Generation R (GENR) study is a population-based birth cohort, established in the Netherlands from children born 2002–06 in the city of Rotterdam and surrounding areas (Kooijman et al., 2016), aimed at examining growth, development and health from fetal life to young adulthood. Data were collected during pregnancy, at birth, and frequently throughout childhood (currently up to age 13). Teacher ratings on behavioral and emotional problems were collected at child mean age 7 years; the response rate was 77%.

The Netherlands Twin Register (NTR) was established in the Netherlands in 1987 and remains ongoing. The register includes adults and young twins who were registered by their parents shortly after birth and who are followed longitudinally to examine, in particular, behavioral development and psychopathology (van Beijsterveldt et al., 2013). Data are collected at ages 1, 2, 3, 5, 7, 9, 10, 12, 14, 16, and 18 years (at which point twins move into the adult twin register). In school year 1999–2000, the NTR began collecting teacher ratings for 7, 10 and 12 year old twins; the average response rate was ~60%.

The Twins Early Development Study (TEDS) was established in the UK from the cohort of twins born 1994-96 (Haworth, Davis, & Plomin, 2013). It is a UK-representative longitudinal twin study aimed at examining language, cognitive, and behavioral development. Data were collected at ages 2, 3, 4, 7, 8, 9, 10, 12, 14, 16, 18, and 21. Teacher ratings on behavioral and emotional problems were collected at ages 7, 9, and 12; response rates were 85%, 76%, 78%, respectively.

### Study questionnaires

The Multidimensional Peer Nomination Inventory (MPNI), used by the FT12 cohort, was originally developed as a tool for rating peers on childhood social behavior, however, it has been adapted and modified to collect ratings from other raters, including teachers (Pulkkinen, Kaprio, & Rose, 1999). It is a 37-item questionnaire with the 6 subscales used here being: aggression (6 items), depression (5 items), hyperactivity–impulsivity (7 items), inattention (4 items), prosocial behavior (12 items), and social anxiety (2 items). Although further subscale scoring was not performed, the MPNI internal structure based on factor analysis indicates aggression, hyperactivity–impulsivity and inattention subscales as part of a behavioral problem/externalizing subscale, and depression and social anxiety subscales as part of an emotional problems/internalizing subscale. Each item on the teacher rating questionnaire has four response choices (from ‘not observed in child’ to ‘clearly observed’). Response choices are scored 0–3, and subscales are formed by taking the mean of all items in the subscale (no missing values were allowed).

The Strengths and Difficulties Questionnaire (SDQ), used by the TEDS cohort, is a 25- item questionnaire that measures common childhood mental health problems (Goodman, 1997). The five subscales of the SDQ are conduct problems (5 items), emotional problems (5 items), hyperactivity (5 items), peer problems (5 items), and prosocial (5 items). The SDQ recognizes conduct problems and hyperactivity subscales as part of an externalizing subscale, and emotional and peer problems subscales as part of an internalizing subscale. For this study, we use ‘conduct problems’ as a proxy for aggressive behavior, and ‘emotional problems’ as a proxy for anxiety problems, as in Bartels et al. (2018). Each item on the questionnaire has three response choices: ‘not true’, ‘somewhat true’, and ‘certainly true’. Response choices range from 0–2 and are coded so that higher scores represent greater risk for the behavior attribute, and subscale scores are derived as a scaled mean score when at least 3 of the 5 items in a subscale are non-missing.

The Teacher Report Form (TRF), part of the Achenbach System of Empirical-Based Assessment’s (ASEBA), was collected by the GENR and NTR cohorts and is a 112-item questionnaire that measures childhood behavioral and emotional problems (Achenbach, 1991). The 8 syndrome subscales used here are aggressive behavior (20 items), anxious/depressed (16 items), attention problems (26 items), rule-breaking behavior (12 items), social problems (11 items), somatic complaints (9 items), thought problems (1 item), and withdrawn/depressed (8 items). The TRF recognizes aggressive behavior and rule-breaking behavior as externalizing problems, and anxious/depressed, withdrawn/depressed and somatic complaints as internalizing problems. In this paper, the attention problems subscale will grouped under externalizing. Each item on the questionnaire has three response choices: ‘not true’, ‘somewhat or sometimes true’, and ‘very true or often true’. Response choices are scored 0–2, and items are summed to created sum scores of individual subscales. A small number of missing values per subscale (depending on the total number of items per subscale), was allowed by replacing the missing values with the mean item score of a subscale.

### Analyses

Data from the FT12, NTR, and TEDS cohorts were analyzed at one site (by AMW) in Stata version 15 (Stata Corporation, College Station, TX, USA), while data from GENR were analyzed locally (by KB) using R version 3.3.2. The analyses followed a standard operating procedure (2018) to ensure uniform data handling and analysis.

First, means, standard deviations (SD) and standard errors (SE) were obtained for all subscales for each age and cohort, separately by gender. T-tests were performed to determine if gender differences existed (p-value <0.05 was considered significant), and effect sizes were calculated as Cohen’s *d* values (with positive values indicating boys have larger mean levels than girls and vice versa). Effect sizes are emphasized over statistical significance since our sample sizes are large and thus significance is expected for many relationships. Next, Pearson correlations between the subscales were computed for each age and cohort, separately by gender.

To formally test gender interactions, as well as to assess the effect size (betas) and amount of variance (R2 values) in aggression explained by co-occurring behaviors together with age, separated by gender, we ran linear regression models. Before modeling, the subscale scores were standardized (mean=0, SD=1) to allow for comparisons of the same questionnaire across different ages or cohorts where possible. Initial modeling included aggressive behavior as the dependent variable and one subscale score as the independent variable, adjusting for age at time of data collection. There were three types of subscale scores in the initial models: 1) the externalizing problems subscales with the highest correlation with aggressive behavior; 2) the internalizing problems subscales with the highest correlation with aggressive behavior; and 3) the prosocial subscale (if available). For each model, data from boys and girls were modeled separately, after testing for gender interactions. Lastly, to examine multiple co-occuring behaviors with aggression, regression modeling was performed with aggressive behavior as the dependent variable and the two (or three, if available) subscale measures from the initial models simultaneously modeled, modeled separately for gender after testing for gender interactions.

## Results

We examined 39,936 teacher ratings on children: 3627 observations from FT12, 4512 observations from GENR, 18,569 observations from NTR, and 13,228 observations from TEDS. There were 17,267 observations of 7-year-olds (49.3% boys), 2762 of 9-year-olds (46.9% boys), 6582 of 10-year-olds (49.6% boys), 11,884 of 12-year-olds (48.4% boys), and 1441 of 14-year-olds (48.4% boys). An interactive summary of all results can be found at (http://www.action-euproject.eu/TeacherRatingsChildAggression).

### Mean Scores

Across all cohorts, mean levels were higher for boys than girls in all externalizing subscale scores, with Cohen’s *d* values ranging 0.31–0.69 (Figure 1 a-c, Online Resource 2).

**Fig. 1 (a-c).**
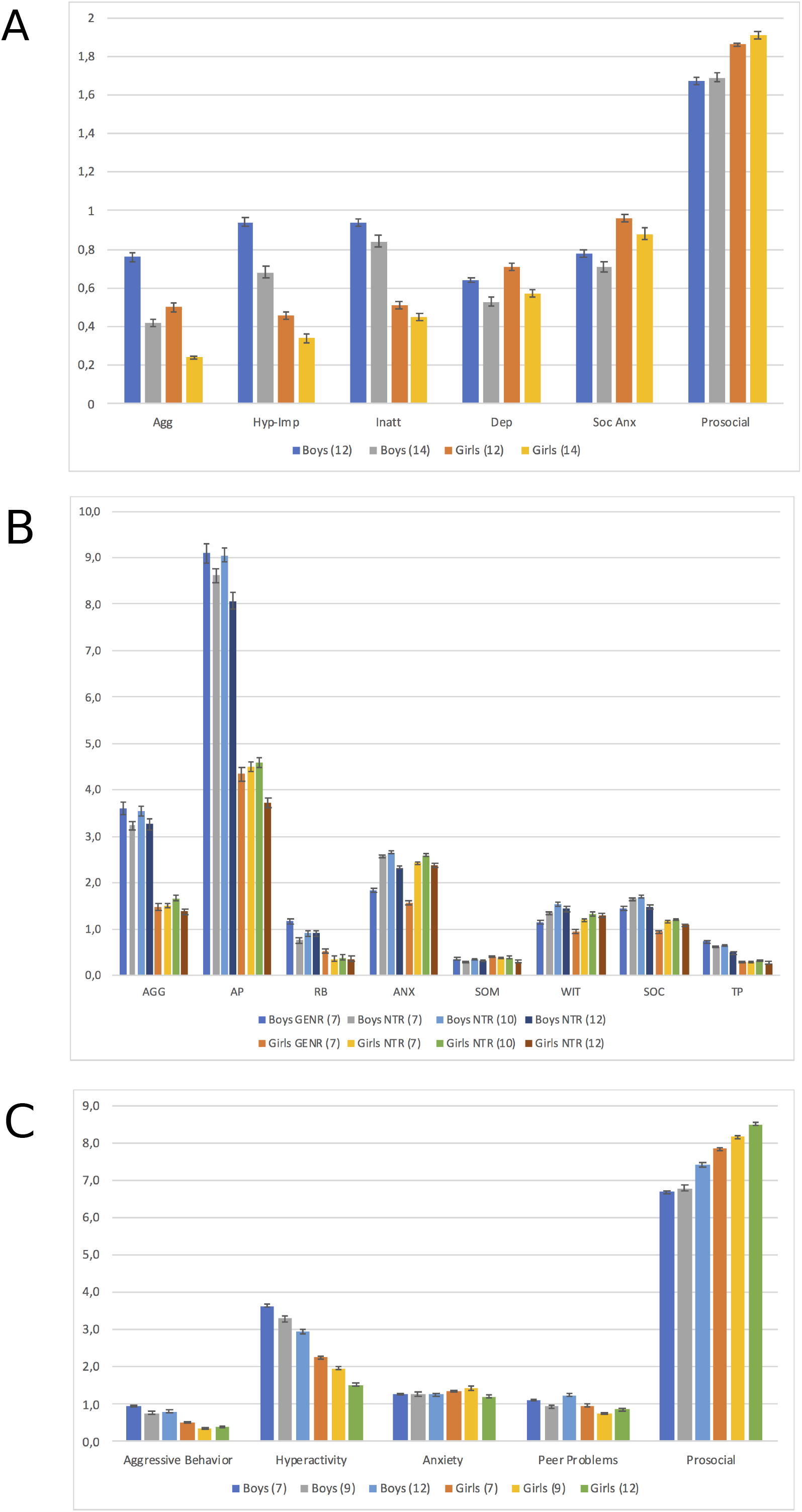
Means and standard errors (SEs) of behavioral scales, separated by gender, age, cohort and behavioral questionnaire **a**. MPNI questionnaire (FT12); Agg=aggression, Dep=depression, Hyp-Imp=hyperactivity- impulsivity, Inatt=inattention, Soc Anx=social anxiety **b**. TRF questionnaire (GENR and NTR); AGG=aggression, ANX=anxious/depressed, AP=attention problems, RB=rule-breaking, SOC=social problems, SOM=somatic problems, TP=thought problems, WIT=withdrawn/depressed **c**. SDQ questionnaire (TEDS)

Results regarding internalizing problems were somewhat cohort dependent, with smaller gender effect sizes than for externalizing problems and prosocial behaviors (Figure 1 a-c, Online Resource 2). In FT12, girls had slightly higher levels of internalizing problems compared to boys (Cohen’s *d* ranged from −0.09 to −0.23). In GENR, NTR and TEDS, for all internalizing problems, boys and girls generally had similar levels (Cohen’s *d* ranged from −0.09 to 0.12).

For all cohorts, mean differences between boys and girls were found in all social behaviors, with girls having higher prosocial scores (Cohen’s *d* range: −0.37–(-)0.63), and boys having slightly more social/peer problems (Cohen’s *d* range: 0.10–0.24).

### Co-occurrence of aggression and other behaviors and problems

In general, correlation patterns of aggressive behavior with other co-occurring behaviors were similar between genders and across cohorts and ages (Figure 2 a-c, Online Resource 3).

**Fig. 2 (a-c).**
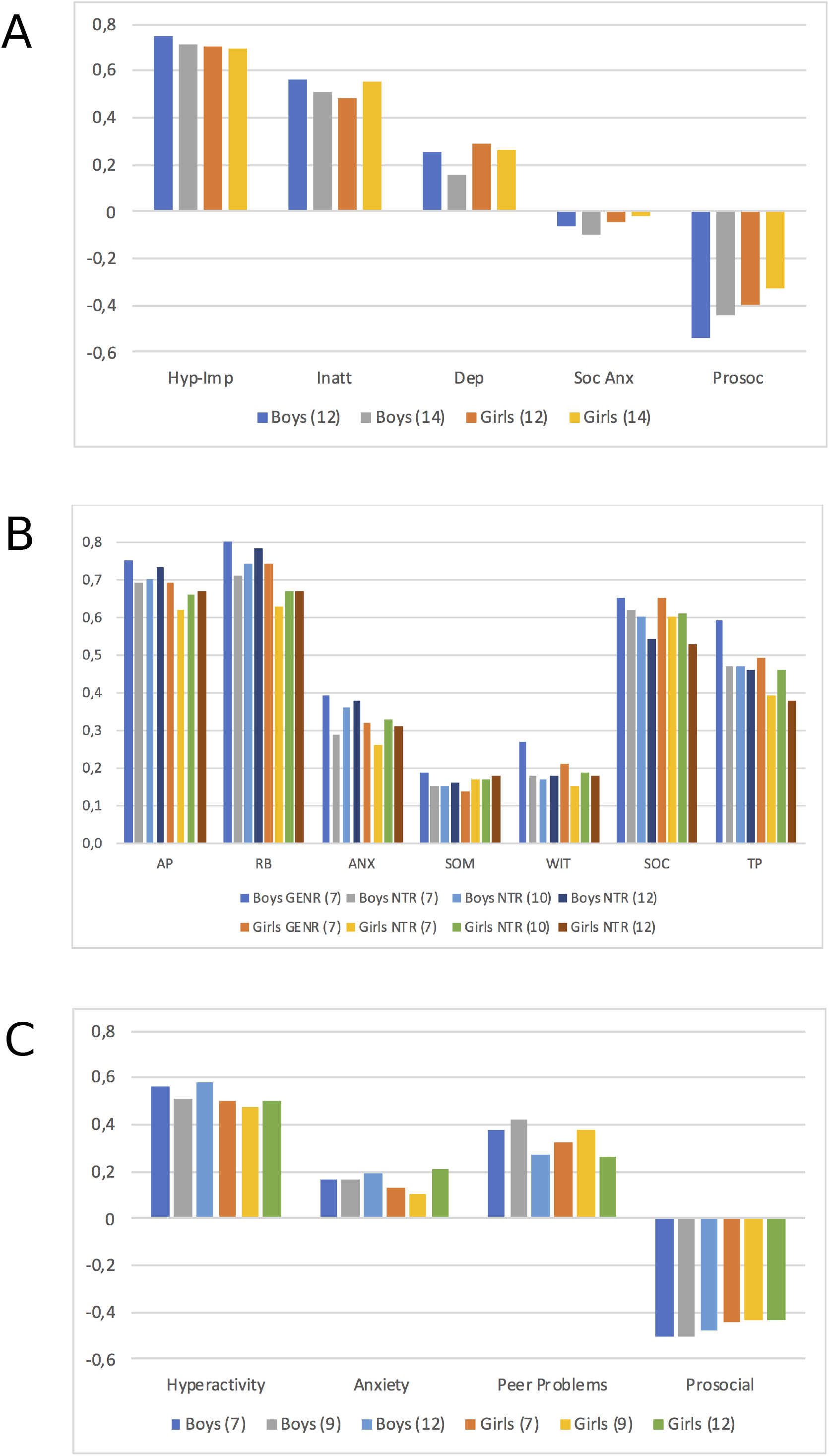
Pearson correlations between aggression and other behavioral/emotional problems, separated by gender, age, cohort and behavioral questionnaire **a**. MPNI questionnaire (FT12); Dep=depression, Hyp-Imp=hyperactivity-impulsivity, Inatt=inattention, Soc Anx=social anxiety **b**. TRF questionnaire (GENR and NTR); AGG=aggression, ANX=anxious/depressed, AP=attention problems, RB=rule-breaking, SOC=social problems, SOM=somatic problems, TP=thought problems, WIT=withdrawn/depressed **c**. SDQ questionnaire (TEDS)

The strength of the correlations of aggressive behavior with other externalizing problems was substantial (Figure 2 a-c, Online Resource 3). Correlations between aggressive behavior and hyperactivity/attention problems ranged 0.51–0.75 for boys and 0.47–0.70 for girls. Correlations of aggressive behavior and rule-breaking behavior (from GENR and NTR) ranged 0.71–0.80 for boys and 0.63–0.74 for girls.

Correlations of aggressive behavior and internalizing problems ranged from small to moderate (Figure 2 a-c, Online Resource 3). Regarding depressive symptoms, correlations with aggressive behavior ranged 0.16–0.27 for boys and 0.15–0.29 for girls. Correlations of general anxiety problems with aggressive behavior (from GENR, NTR, and TEDS) ranged 0.17–0.39 in boys and 0.10–0.33 in girls, whereas social anxiety and aggressive behavior (from FT12) correlations ranged from −0.02 to −0.10 for boys and girls.

Correlations of aggressive behavior and social behavior were moderate (Figure 2 a-c, Online Resource 3). Prosocial and aggressive behavior correlations (from FT12 and TEDS) ranged from −0.44 to −0.54 for boys and −0.33 to −0.44 for girls. Aggressive behavior and social/peer problems (from GENR, NTR, and TEDS) ranged 0.27–0.65 for boys and 0.26–0.65 for girls.

### Supplemental Analyses (Modeling)

In linear regression modeling (i.e., aggressive behavior as the dependent and one subscale as the independent variable), gender was significantly associated with aggression. Additionally, all GENR and NTR and nearly all TEDS models indicated significant gender interaction terms (Online Resource 4). In contrast, half of the FT12 models did not indicate significant gender interaction. Regarding model R_2_ values (% of variance in dependent variable explained by independent variables), externalizing models were consistently largest (boy range: 0.26–0.63; girl range: 0.22–0.54). For internalizing models, R2 values were small (boy range: 0.03–0.15; girl range: 0.01–0.11). For prosocial models, R2 values ranged 0.11–0.29 (boy range: 0.19–0.29; girl range: 0.11–0.19).

In further regression modeling (i.e., multiple subscales included simultaneously as independent variables), gender interactions were generally significant. R_2_ values generally did not differ from those in externalizing-only models (Online Resource 5). However, independent variables in these analyses were generally all significant, though with smaller effect sizes compared to initial models.

## Discussion

This study provides a reference panel of average levels and co-occurence of aggressive behavior and other behaviors and emotional problems for schoolchildren ages 7–14 in the European school setting. Although it is commonly assumed that aggressive behavior is associated with these other problems and behaviors, this is one of the few studies to look at large, population-based samples across multiple countries. Results indicate that patterns are quite similar across the different cohorts of Finland, the Netherlands, and the UK. We show that, as expected, the levels of aggressive behavior are statistically significantly different by gender, however, the effect sizes are only moderate. We also see that aggression often co-occurs with other behavioral and emotional difficulties, in both boys and girls, and that these correlations are quite similar between the genders. Regression modeling indicated that much of the variation (R_2_) in aggressive behavior levels was explained by other co-occurring externalizing problems, though prosocial models also had rather large R_2_ values in initial individual models. Modeling multiple co-occurring behaviors simultaneously indicated that children with aggressive behavior often have not only one co-occurring problem behavior, but multiple co-occurring problems (including low prosocial skills).

Regarding co-occurence of aggressive behavior with other externalizing problems, we see remarkable similarities in the gender patterns. Although boys had the expected higher levels and associations of aggression and other externalizing problems compared to girls, correlations between aggression and other externalizing behaviors were very similar (r differences ≤0.1 between boys and girls). In this respect, we can point to another study on the NTR cohort regarding gender differences in ADHD diagnosis and comorbidity using teacher ratings (Derks, Hudziak, & Boomsma, 2007) in which boys and girls had similar comorbidity profiles and school impairment. However, girls were less likely than boys to be identified by teachers as disruptive and referred for treatment. Similarly, conduct disorder and oppositional defiant disorder have both been much more likely to be attributed to boys than girls among teacher ratings, compared to parent ratings (Maughan, Rowe, Messer, Goodman, & Meltzer, 2004; Offord et al., 1996). While keeping situational aggressive behavior and discrepancies between raters in mind (Achenbach et al., 1987; De Los Reyes & Kazdin, 2005; Verhulst et al., 1994), we also need to consider that girls may not be as readily identified as problematic regarding aggression by teachers as boys, although their patterns of comorbid problems and need for treatment may be similar.

While the relationships between aggressive behavior and co-occurring externalizing problems are well-established, the co-occurrence with internalizing problems are less well described. Generally, there is moderate correlation between aggressive and depressive symptoms (van den Oord, Verhulst, & Boomsma, 1996), which we saw in our cohorts as well. However, the relationship with anxiety is less well characterized. The range of anxiety correlations were weak or moderate (positive for general anxiety measure, negative for social anxiety), and wider ranging than the depressive symptoms and aggressive behavior correlations. These mixed results regarding aggressive behavior and internalizing problems could partially be explained by August et al. (1996) who found that, among schoolchildren, anxiety/mood disorders most often co-occur with multiple externalizing disorders (instead of, e.g., only conduct disorder). We also saw, in the multiple co-occuring behaviors models, that internalizing problems tend to co-exist alongside both aggressive behavior and other co-occuring externalizing behaviors. One note of striking similarity regarding internalizing problems and aggressive behavior was between boys and girls (r differences ≤0.08 between boys and girls). Future studies should further clarify these similarities and differences in aggressive behavior and internalizing problems, which are perhaps less often considered in the school setting than the more overt and administratively problematic externalizing behaviors.

Prosocial behavior was negatively associated with aggressive behavior, as has been observed by others (Eron & Huesmann, 1984), with moderate strength. In regression-based models, prosocial behavior explained much of the variance in aggressive behavior, but the effect of prosocial behavior attenuated substantially (though remained significant) when externalizing behavior was added to the model. The relationships are thus complex, aggressive students are often struggling not only with other externalizing problem behaviors but also a lack of social skills. It is of interest to note that Kokko and Pulkkinen (2000) observed that prosocial behavior is protective in aggressive children against future unemployment. Additionally, Hämäläinen and Pulkkinen (1996) have shown that aggressiveness without other co-occurring problems did not predict future criminality, while those with an accumulation of behavioral problems (including aggressive behavior and poor prosocial skills) predicted future criminality best. Interestingly, they also showed that low prosocial behavior alone predicted future criminality. Furthermore, although statistically significant, we saw that prosocial–aggressive behavior correlations by gender were <0.15 different between boys and girls, and the differences between genders in TEDS correlation values were half of those in FT12. This may be related to the severity of the aggression measure, since the SDQ (used in TEDS) captures conduct problems, which are more serious than the general aggression captured by the MPNI (used in FT12), thus suggesting that, especially when comparing the highest levels of aggression with levels of prosocial behavior, there are no gender differences.

In our study protocol, each age from each cohort represents a cross-sectional snapshot of behavioral and emotional problems, with some children captured in more than one age category. Because all data are drawn in large numbers and in a population-based manner, these data do reflect developmental stages well. Over the ages (7–14 years), we can see that patterns do not generally differ by gender. For all cohorts, the differences in aggressive–co-occurring behavior correlations between all ages were <0.11. In longitudinal settings, the general trend is that population levels of aggressive behavior diminish as age increases, although there tends to be stability in rank order across certain individuals (those with high aggression and those of lower socioeconomic status) (Odgers et al., 2008; Pulkkinen, 2017).

Although detailed comparison between cohorts regarding behavioral questionnaires, countries and school systems are outside the scope of this paper (however, see (Hendriks et al., 2019) and Online Resource 1, for information on these aspects in the cohorts and countries), it is noteworthy that we see similar patterns across cohorts (questionnaires), since the cohorts represent different European countries and the questionnaires were developed for different purposes. Indeed, future studies should consider validating their findings across other countries due to the robustness of the results across countries and questionnaires. Furthermore, both the ASEBA system (of which the TRF is a part) and the SDQ were assessed by Achenbach et al. (2008) and generally found to produce comparable results across countries, with more variation found within populations than between. Moreover, we have found that the negative relationship between aggression and academic performance was consistent and generally similar in ACTION cohorts with different aggression questionnaires (Vuoksimaa, 2020 in press).

One final comparison can be made to a parallel analysis among the same ACTION cohorts, using parental and self ratings ((Bartels et al., 2018); interactive results also available at http://www.action-euproject.eu/ComorbidityChildAggression). Overall, teacher-based correlations of aggressive behavior with co-occuring behaviors are higher for externalizing problems and lower for internalizing problems and prosocial behaviors than parental ratings at similar ages.

Despite the nearly 40,000 teacher-rated observations of behavioral and emotional problems among school children ages 7–14 collected across 3 European countries, there are limitations to consider in this study. The data come from three higher income countries in Europe, and it is unclear if these patterns would remain in lower income countries or countries of differing cultural backgrounds. For example, a recent publication on data from 63 low- and middle-income countries collected by the World Health Organization’s Global School-based Student Health Survey indicates that gender differences in physical aggression are stronger as a function of greater gender equality in a society (Nivette et al., 2018).

Lastly, three of the four cohorts consist of twins. While twins are born on average prematurely and of lower birthweight, they are generally indistinguishable from singletons later in childhood on multiple traits and conditions. Studies from, for example, FT12 and NTR (de Zeeuw, van Beijsterveldt, de Geus, & Boomsma, 2012; Pulkkinen, Vaalamo, Hietala, Kaprio, & Rose, 2003) have shown that the twins are representative of schoolchildren with respect to both internalizing and externalizing behaviors and educational achievement. Additionally, twins are born in every stratum of society, and twins’ parents and teachers are generally well motivated to take part in research.

We have presented a reference set of teacher-rated childhood aggressive behavior levels and associations with co-occurring behaviors in the school setting. These results indicate that aggressive behavior regularly co-occurs with other externalizing behaviors and a lack of prosocial skills, and moderately co-occurs with internalizing problems. Furthermore, we draw attention to the relative similarities in patterns of associations between aggressive behavior and co-occurring behaviors across genders and participating cohorts using different measures. Teachers are a valuable resource for identifying children in need of specific support and possibly in referral or diagnostic aspects. However, teacher trainings could help to reduce potential gender prejudice regarding problem behaviors and to recognize that those with one problem behavior likely have multiple problem behaviors they are struggling with. Additionally, school interventions for aggression need to be holistic, focusing on broad behavioral and emotional improvement including support to develop prosocial skills, such as the successful “multi-year universal social–emotional learning program” implemented by Greenberg et al. (2010) that showed reduced levels of aggression and increased prosocial skills.

## Compliance with Ethical Standards

All data in the current investigation were collected under protocols that have been approved by the appropriate ethics committees, and studies were performed in accordance with the ethical standards established in the 1964 Declaration of Helsinki and its later amendments. All participants (or their guardians) provided informed consent before participation in their respective cohorts.

All authors declare no conflicts of interest.

## Data Availability

The data that support the findings of this study are not publicly available due to privacy or ethical restrictions. The corresponding author is able to provide analysis code and information on how to access the cohorts and request the data from the appropriate managers of each cohort.

## DECLARATIONS

### Ethics Approval

All data in the current investigation were collected under protocols that have been approved by the appropriate ethics committees, and studies were performed in accordance with the ethical standards established in the 1964 Declaration of Helsinki and its later amendments.

### Consent to Participate

All participants (or their guardians) provided informed consent before participation in their respective cohorts.

### Consent for Publication

No identifying information from participants is used in this study. The general consents provided by participants allows for aggregate results to be published.

### Code Availability

The code used for the study is available upon reasonable request from the corresponding author.

## Acknowledgements

This work is part of the ACTION consortium which is supported by funding from the European Union Seventh Framework Programme (FP7/2007-2013) under grant agreement no. 602768.

FT12: We gratefully acknowledge the ongoing contribution of the participating twin families. Data collection has been supported by the National Institute of Alcohol Abuse and Alcoholism (grants AA-12502, AA-00145, and AA-09203 to RJR) and the Academy of Finland (grants 100499, 205585, 118555, 141054 and 264146 to JK). JK has been supported by the Academy of Finland (grants 308248, 312073). EV has been supported by the Academy Research Fellow funding from the Academy of Finland (grant 314639).

GENR: This work was supported by the Netherlands Organization for Scientific Research (NWO-grant 016.VICI.170.200) to HT, and the Sophia Children’s Hospital Research Foundation (research fellowship grant 921) to KB. Super computing resources were made possible through the NWO Physical Sciences Division (surfsara.nl).

The first phase of the Generation R Study is made possible by financial support from the Erasmus Medical Centre, Rotterdam; the Erasmus University Rotterdam; The Netherlands Organization for Health Research and Development (ZonMw). The authors gratefully acknowledge the contribution of all children and parents, general practitioners, hospitals, midwives, and pharmacies involved in the Generation R Study. The Generation R Study is conducted by the Erasmus Medical Center (Rotterdam) in close collaboration with the School of Law and Faculty of Social Sciences of the Erasmus University Rotterdam; the Municipal Health Service Rotterdam area, Rotterdam; the Rotterdam Homecare Foundation, Rotterdam; and the Stichting Trombosedienst & Artsenlaboratorium Rijnmond, Rotterdam.

NTR: We gratefully acknowledge the ongoing contribution of the participating familes. Data collection in the NTR was funded by the Netherlands Organization for Science (NWO): Twin- family database for behavior genetics and genomics studies (NWO-480-04-004); Gravitation program of the Dutch Ministry of Education, Culture, and Science and the Netherlands Organization for Scientific Research (NWO-024.001.003). ‘Longitudinal data collection from teachers of Dutch twins and their siblings’ (NWO-481-08-011); ‘Twin-family study of individual differences in school achievement’ (NWO-056-32-010); ZonMW “Genetic influences on stability and change in psychopathology from childhood to young adulthood” (NW0-912-10- 020); “Netherlands Twin Registry Repository” (NW0-480-15-001/674). DIB would like to acknowledge the KNAW Academy Professor Award (PAH/6635).

TEDS: We gratefully acknowledge the ongoing contribution of the participants in the Twins Early Development Study (TEDS) and their families. TEDS is supported by a program grant to RP from the UK Medical Research Council (MR/M021475/1).

No funders had any involvement in the conduct of this research, including in the study design, data collection, analysis and interpretation of the data, the writing of the report, and the decision to submit to the article for publication.

